# Analytical and clinical validation of a circulating tumor DNA-based assay for multi-cancer early detection

**DOI:** 10.1101/2023.12.22.23300420

**Authors:** Luu Hong Dang Nguyen, Thi Hue Hanh Nguyen, Van Hoi Le, Vinh Quang Bui, Lan Hieu Nguyen, Nhu Hiep Pham, Thanh Hai Phan, Huu Thinh Nguyen, Van Song Tran, Chi Viet Bui, Van Kha Vo, Pham Thanh Nhan Nguyen, Ha Huu Phuoc Dang, Van Dung Pham, Van Thinh Cao, Ngoc Minh Phan, Ba Linh Tieu, Giang Thi Huong Nguyen, Dac Ho Vo, Trung Hieu Tran, Thanh Dat Nguyen, Van Thien Chi Nguyen, Trong Hieu Nguyen, Vu Uyen Tran, Minh Phong Le, Thi Minh Thu Tran, Minh Nguyen Nguyen, Thi Tuong Vi Van, Anh Nhu Nguyen, Thi Thanh Nguyen, Nhu Nhat Tan Doan, Hoang Tan Nguyen, Phuoc Loc Doan, Le Anh Khoa Huynh, Tien Anh Nguyen, Huu Tam Phuc Nguyen, Y-Thanh Lu, Chi Thuy Tien Cao, Van Tung Nguyen, Thi Le Quyen Le, Thi Lan-Anh Luong, Thi Kim Phuong Doan, Thi Trang Dao, Canh Duy Phan, Thanh Xuan Nguyen, Nguyen Tuong Pham, Bao Toan Nguyen, Thi Thu Thuy Pham, Huu Linh Le, Cong Thanh Truong, Thanh Xuan Jasmine, Minh Chi Le, Van Bau Phan, Quang Binh Truong, Thi Huong Ly Tran, Minh Thien Huynh, Tu Quy Tran, Si Tuan Nguyen, Vu Tran, Van Khanh Tran, Huu Nguyen Nguyen, Duy Sinh Nguyen, Thi Van Phan, Thi Thanh-Thuy Do, Dinh Kiet Truong, Hung Sang Tang, Hoa Giang, Hoai-Nghia Nguyen, Minh-Duy Phan, Le Son Tran

## Abstract

**Background:** The emergence of multi-cancer early detection (MCED) via a single blood test offers promise in enhancing the efficiency of early cancer detection and improving population health. However, the lack of analytical validation and clinical evidence across diverse populations has hindered their adoption in clinical practice. To address these challenges, we undertook a comprehensive analytical and clinical validation for our MCED test, SPOT-MAS (Screening for the Presence Of Tumor by DNA Methylation And Size).

**Methods:** The analytical validation was conducted on a retrospective cohort of 290 healthy and 461 cancer-confirmed individuals to establish the limit of detection, repeatability and reproducibility of test results and assess the impact of potential interferents on test performance. To validate the performance of SPOT-MAS test in clinical settings, we launched a multi-center prospective trial, named K-DETEK, of 9,057 asymptomatic participants in Vietnam.

**Findings:** For analytical validation, SPOT-MAS could detect at least 50% of cancer samples at a specificity of 98% if the samples have tumor fraction 0.049 (95% CI: 0.043-0.059). The results were consistently reproduced for both intra- and inter-batch analysis. Moreover, our test remained robust at hemoglobin contamination of 500 mg/dl and genomic DNA contamination of up to 100%. In the clinical trial, our assay achieved a positive predictive value of 58.14% (95%CI: 43.33-71.62) with 84.00% (95%CI: 65.35-93.60) accuracy in predicting tumor location, a negative predictive value of 99.92% (95%CI: 99.84-99.96), an overall sensitivity of 78.13% (95% CI: 61.25-88.98) and a specificity of 99.80% (95% CI: 99.68-99.87).

**Interpretation:** To our knowledge, this is the first and largest prospective validation study in Asia supporting the utility of SPOT-MAS as a multi-cancer blood test for early detection in a limited-resource country, where a nationwide cancer screening program is urgently needed but currently not available.

**Funding:** Gene Solutions

## BACKGROUND

Cancer is the second leading cause of death and continues to have a significant impact on global mortality rates. The prevalence of cancer and its associated mortality has put a strain on healthcare systems worldwide^1^. Detection in advanced stages of the disease has also increased the burden of cancer^2^. In an effort to combat this, conventional cancer screening methods, such as those recommended by the United States Preventive Services Task Force, have shown promise in increasing overall survival rates, improving treatment efficiency, and reducing long-term medical costs^3–5^. However, certain existing screening methods, particularly colonoscopy or cervical cytology tests, are invasive and have low accessibility^6,7^. Moreover, they focus solely on detecting a single cancer type, leading to a high cumulative false positive rate when performed sequentially^8^. One of the most promising advancements in cancer detection is the development of non-invasive multi-cancer early detection (MCED) tests^9^. MCED tests, using blood-based liquid biopsy (LB) approaches, have the potential to revolutionize cancer screening by enabling early detection of multiple types of cancer through a simple blood draw^10^. LB assays detect specific cancer-related biomolecules including circulating tumour cells (CTC), cell free DNA (cfDNA), circulating tumor DNA (ctDNA), circulating free RNA (cfRNA) and exosomes^11^. Of these, ctDNA released into the circulation when tumor cells undergo apoptotic and necrotic cell death processes has been extensively studied due to its tissue- and cancer-type specificity^12–14^. Recently, the landscape of MCED tests based on detecting methylation changes in cfDNA has evolved. The OverC test (Burning Rock) or Galleri test (Grail) can detect multiple cancer types simultaneously with high performance by interrogating methylation changes in cfDNA^15,16^. The Galleri test has been clinically validated in asymptomatic (PATHFINDER study) and symptomatic population (SYMPLIFY study). The Galleri test was shown to exhibit a PPV of 38% for detecting cancer and the tumor of origin (TOO) prediction with an accuracy of 97% in an asymptomatic population^15^. Moreover, for individuals with malignancy-related symptoms, the SYMPLIFY trial in England and Wales demonstrated a particularly high PPV of 75.5% and NPV of 97.6%^16^.

Despite promising results, MCED methods demonstrated low sensitivity for detecting certain cancers (e.g., breast cancer) and early-stage tumors owing to low amount and high heterogeneity of ctDNA^10,17,18^. To improve the detection sensitivity of ctDNA, MCED screening methods tend to use high-depth sequencing, making it economically impractical for population-wide screening^10,19^. To address these limitations, we have recently developed a multimodal method, known as Screening for the Presence Of Tumor by Methylation And Size (SPOT-MAS), to simultaneously detect five common types of cancer, including liver, breast, colorectal, gastric, and lung cancer, and predict the tissue origin of cancer signal^17,20,21^. By integrating cost-effective shallow sequencing and advanced machine learning, SPOT-MAS was trained and validated on a large cohort of 2,288 participants, including 738 nonmetastatic cancer patients and 1,550 healthy controls, achieving a sensitivity of 72.4% at 97.0% specificity and an accuracy of 70.0% for tumor-of-origin (TOO) prediction^21^. We further assessed the performance of SPOT-MAS in an interim 6-month study, named K-DETEK, which involved 2,795 participants at 13 clinical centers and one research institue in Vietnam^22^. In this study, we conducted the analytical validation of the SPOT-MAS assay on a retrospective cohort of 290 healthy and 461 cancer-confirmed individuals to determine the limit of detection and investigate the impact of technical factors on the robustness of our assay (**Figure 1**). Moreover, we prospectively recruited 9,057 asymptomatic participants at 75 clinical centers and one research institute in Vietnam to comprehensively evaluate the clinical performance of SPOT-MAS and demonstrate its clinical utility in the early detection of multiple types of cancer (**Figure 1**). The clinical applicability of SPOT-MAS is measured by positive predictive value (PPV), negative predictive value (NPV), sensitivity, specificity and accuracy of TOO prediction.

**Figure 1.**
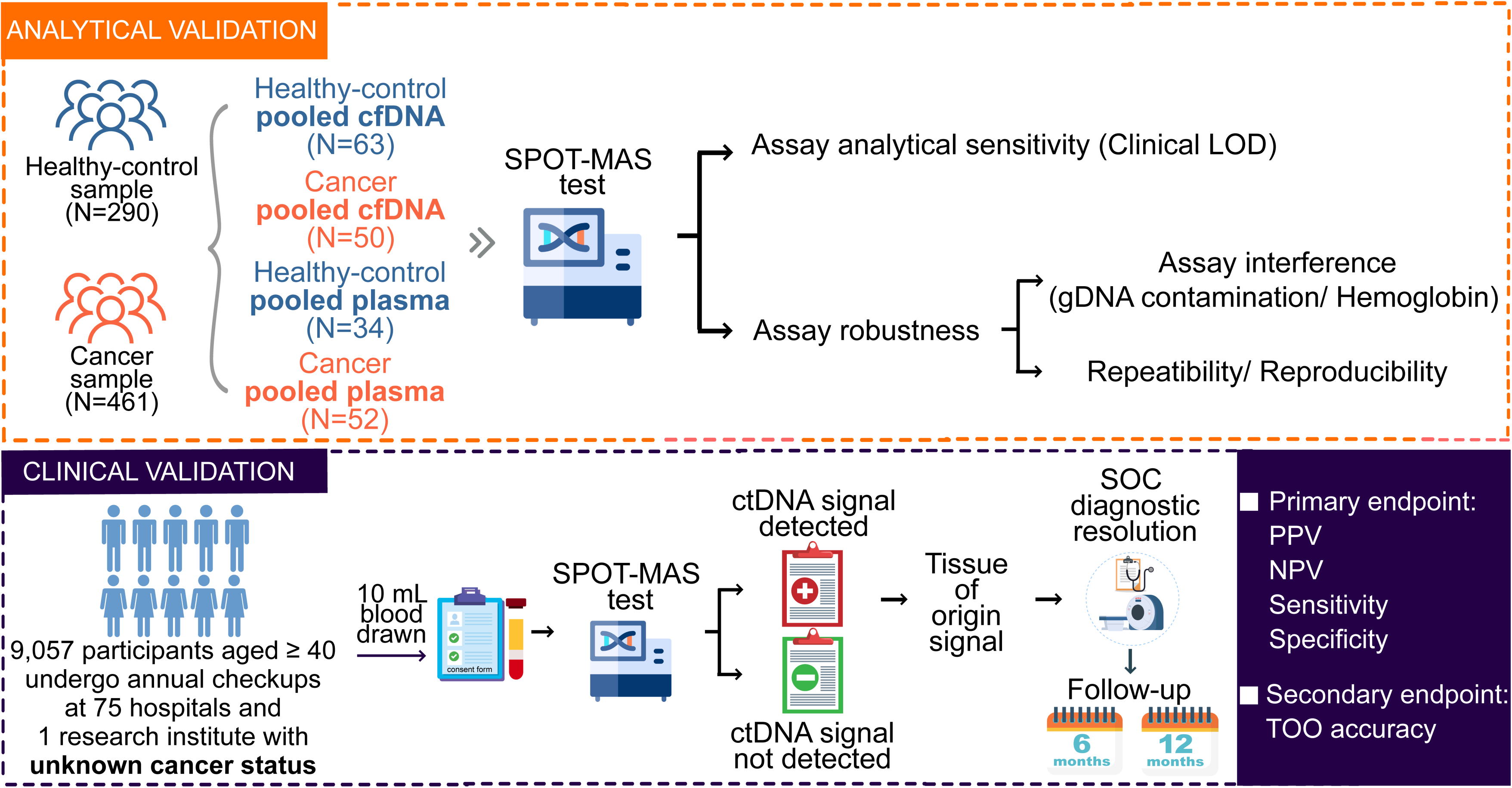
Overview of analytical and clinical validation. For analytical validation, two investigations were conducted: i) Determination of assay analytical sensitivity (or clinical limit of detection); and ii) Assessment of assay robustness by evaluating potential interferences (genomic DNA and hemoglobin) and assay repeatability and reproducibility. For clinical validation, K-DETEK is a prospective study recruiting 9,057 asymptomatic participants aged 40 or older from 75 hospitals and one research institute in Vietnam. Plasma cfDNA was extracted from 10 ml of blood from eligible participants, and the cfDNA extraction was analyzed using the SPOT-MAS assay. SPOT-MAS provides two test results: “ctDNA signal detected” or “ ctDNA signal not detected” along with the predicted tumor of origin. Participants with a ctDNA signal detected results were consulted by physicians and underwent confirmation through diagnostic imaging tests or based on the TOO probability values. All participants were followed up after 6 and 12 months to obtain information on possible cancer diagnoses.

## METRIALS AND METHODS

### Analytical validation

Analytical validation was undertaken to establish the clinical limit of detection (LOD) and assess the robustness of the SPOT-MAS assay (**Figure 1**).

#### Analytical sample preparation

For analytical analysis, samples were collected from 290 healthy and 461 diagnostic-confirmed individuals for each type of cancer from our previous retrospective study^21^. The demographic details of the samples used for analytical validation are listed in Table S1. Healthy samples were collected from participants who were confirmed cancer-free at the time of enrollment and followed up for three years after enrollment to ensure that they did not develop cancer. For each cancer type, including breast, colorectal, gastric, liver, and lung cancer, the cancer diagnosis and staging was confirmed by pathology and imaging analysis based on the American Joint Committee on Cancer (AJCC) Staging Manual (version 8). To obtain sufficient materials for analytical validation, we generated pooled cfDNA or pooled plasma samples. Specifically, 63 healthy and 50 cancer pooled cfDNA samples were generated by mixing cfDNA isolated from different healthy subjects or cancer patients diagnosed with the same cancer type and used for LOD determination (**Table S1**). Moreover, a total of 34 healthy and 52 cancer pooled plasma samples were generated by mixing 1 ml of plasma isolated from healthy subjects or patients diagnosed with the same cancer type and used for assay robustness assessment (**Table S1**).

### Tumor fraction estimation and establishing clinical limit of detection

Sixty-three pooled cfDNA samples from healthy individuals and 50 pooled plasma samples from cancer patients, including breast cancer (n=13), colorectal cancer (n=12), liver cancer (n=10), lung cancer (n=9), and gastric cancer (n=6), were subjected to the SPOT-MAS assay. The tumor fraction (TF) of cancer cfDNA samples was determined by the ichorCNA tool, which predicts segments of somatic copy number alterations (CNAs) and estimates TF while accounting for subclonality and tumor ploidy^23^. Then, the cancer cfDNA samples with known TF were spiked into the healthy cfDNA samples at various levels of 0.5%, 1%, 5%, 15%, 25%, 50%, and 100% of the neat samples with estimated TF. The LOD of SPOT-MAS for each type of cancer was defined as the TF levels at which the probability of detecting a cancer signal was at least 50%, while maintaining at least 98% specificity, as previously described by Jamshidi et al.^24^.

#### Evaluation of assay robustness

To determine the effect of potential substances that could interfere with the performance of SPOT-MAS assay, 20 cancer and 10 healthy-control pooled plasma samples were spiked with genomic DNA or hemoglobin. Each pooled plasma sample was spiked with genomic DNA (0, 50, 100, 150, 200%) or hemoglobin (0, 100, 500, 1000 and 2000 mg/dL).

To determine the ability to reproduce and variability of SPOT-MAS assay, a total of 56 pooled plasma samples including 32 cancer and 24 healthy-control were used. The concordance rate was analyzed within runs (intra-batch) and between runs (inter-batch) with different lots of reagents and operators. For the analysis of intra-batch variations, each sample was performed in triplicate (denoted by A, B and C). For the analysis of inter-batch, each sample was divided into three independent batches (denoted by 1, 2, and 3). Samples were subjected to SPOT-MAS assay and the consistency was evaluated based on the prediction results of SPOT-MAS.

### Clinical validation

#### Study design

The K-DETEK study is a prospective investigation aimed at evaluating the performance of the multi-cancer early detection test, SPOT-MAS, in detecting asymptomatic cancer within a screening-relevant population. The study recruited 9,057 participants having follow-up visits for chronic conditions or undergoing annual health check-ups across 75 hospitals and one research institute in Vietnam from April 2022 to April 2023. The study was registered with the U.S. National Institutes of Health (ClinicalTrials.gov identifier: NCT05227261). The institutional ethics and scientific committee of the University of Medicine and Pharmacy, Ho Chi Minh City, Vietnam, reviewed and approved this study (approval number: 192/HĐĐĐ-ĐHYD). All participants provided written informed consent prior to participating in this study. Participants were eligible for K-DETEK study if they were aged 40 years or older, willing to return for required follow-up visits at 6 months and 12 months, had neither clinical suspicion of cancer nor history of confirmed cancer, had no history of blood transfusion or bone marrow transplantation in 3 months prior to recruitment time and had no clinical manifestations of pregnant. All the clinical characteristics of participants are de-identified and listed in **Table S2**.

#### Laboratory Workflow

Blood samples of 10 ml were collected in Streck cfDNA tubes and transported to the central laboratory for plasma cfDNA extraction. The median time from blood collection to plasma isolation was 2 days, ranging from 0 to 5 days. Subsequently, the isolated cfDNA underwent the SPOT-MAS assay, following previously described protocols^21^. The multiple features of cfDNA, including 450 target regions’ methylation, genome-wide methylation profiles (global methylation density of 2734 1Mb-bins on 22 chromosomes), fragment length, DNA copy number of 588 5Mb-bins on 22 chromosomes and end motifs, were simultaneously analyzed by SPOT-MAS workflow^21^. Using machine learning algorithms, the model returned the probability scores of ctDNA signal detection (SPOT-MAS score) and the tumor of origin (TOO) for those with ctDNA signal detected (**Figure 1**).

#### Informing participants of test results

The SPOT-MAS test results were returned to the study participants within a 30-day period following blood collection. Time to diagnostic resolution for each participant was calculated as the duration in days between the availability of test results to the ordering physicians and the date of diagnostic resolution, as determined by the ordering physicians. SPOT-MAS provides two types of test results: “ctDNA signal not detected” (negative) or “ctDNA signal detected” (positive), with up to two prediction results for TOO.

The participants with a positive result were consulted by physicians to undertake diagnostic imaging tests according to the TOO probability values across five types of cancer: breast, liver, lung, colorectal and gastric. TOO signals that are not covered by SPOT-MAS test were reported as “other cancer”. Cancer diagnoses were reported according to The National Comprehensive Cancer Network (NCCN) for cancer screening. The diagnosis tests are listed in **Table S3**. Participants with “other cancer” status were recommended for a whole-body CT scan for cancer screening. Participants with a negative result were informed about their lack of risk for the five cancer types covered by SPOT-MAS test. All participants were followed up after 6 months and 12 months to obtain information on possible cancer diagnosis.

The performance of SPOT-MAS assay was determined, including true positive rate, false positive rate, positive predictive value (PPV), negative predictive value (NPV), sensitivity and specificity for cancer signal detection (%). The overall prediction accuracy (%) of TOO was also assessed.

#### Participant demographic and statistical analysis

The demographic information of all participants was listed in **Table S2**. Participants in the high-risk group were identified based on factors such as heavy smoking, alcohol consumption, hepatitis B/C infection, type 2 diabetes, or having first-degree relatives (FDR) diagnosed with two cancer types at an age younger than 45 or being identified as mutant carriers. The remaining participants were classified into the moderate-risk group.

All statistical analyses were performed by using R (4.3.2) with standard data analysis packages and the ggplot2 package for visualization. Confidence intervals were analyzed by Wilson method using R (4.3.2)^25^. Sigmoid curves with probit regression were used for LOD analysis using MedCalc software version 22.001 (MedCals Software, Ostend, Belgium; https://www.medcalc.org; 2020). Correlation analysis between tumor fractions estimated by ichorCNA and percentage of spike-in were measured by Pearson coefficient R.

## RESULTS

### The limit of detection of SPOT-MAS assay

Limit of detection (LOD) is a critical parameter to assess the analytical sensitivity of an assay. To obtain sufficient materials for the evaluation of LOD, we first generated 63 healthy and 50 cancer cfDNA samples (13 breast cancer, 12 colorectal cancer, 10 liver cancer, 9 lung cancer and 6 gastric cancer) by pooling cfDNA samples of healthy individuals or patients diagnosed with the same cancer type as described in the method section (**Figure 2A**). Subsequently, we determined tumor fraction (TF) of these pooled cfDNA samples by using the ichorCNA tool that was previously developed by Viktor et al.^23^ The ichorCNA tool was originally developed to quantify TF in low-pass non-bisulfite genome-wide sequencing reads. To demonstrate the feasibility of using ichorCNA for our bisulfite-converted cfDNA, we randomly selected cfDNA from 6 cancer and 4 healthy plasma samples to perform a comparison of TF levels between bisulfite and non-bisulfite sequencing results. We observed an excellent correlation between tumor fraction of bisulfite-treated and non-bisulfite-treated fragments calculated by ichorCNA for all 10 samples (Pearson correlation, R = 0.99, p= 1.7.10^-11^, **Figure S1**), indicating that the ichorCNA tool can be applied to bisulfite-converted sequencing cfDNA samples. After quantifying TF of our pooled cfDNA samples, we spiked the cancer pooled cfDNA with known TF into the healthy pooled cfDNA samples at different fractions of cancer cfDNA (0.5, 1, 5, 15, 25, 50 and 100%). As a result, for each cancer type we have from 6 to 13 cancer pooled cfDNA samples with known TF and we obtained seven spike-in cfDNA samples per cancer pooled cfDNA sample, giving us a total of 350 spike-in samples for the 50 cancer cfDNA samples (**Table S4**). We observed significant positive correlations between TF levels and spike-in concentrations in most cancer types, with the highest correlation coefficient in liver cancer (R=0.84, p< 2.2.10^-6^, **Figure 2B**), followed by colorectal cancer (R= 0.82, p< 2.2.10^-6^, **Figure 2B**), lung cancer (R= 0.72, p< 2.2.10^-6^, **Figure 2B**), gastric cancer (R= 0.57, p=1.4.10^-10^, **Figure 2B**) and breast cancer (R=0.47, p=6.5.10^-10^, **Figure 2B**). Thus, these results demonstrated the potential of the ichorCNA based approach in quantifying tumor fraction in our spike-in samples.

**Figure 2.**
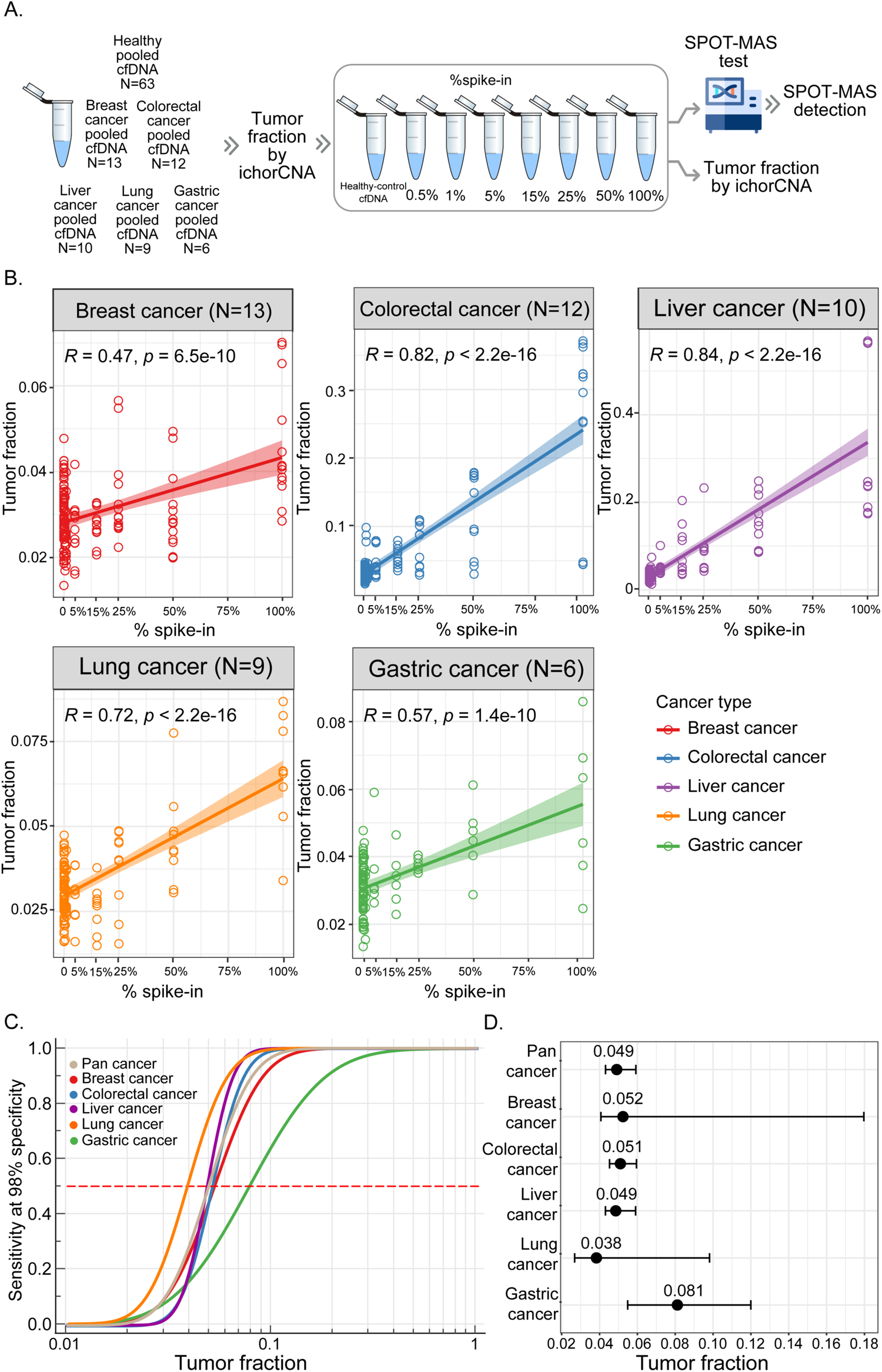
Determination of limit of detection. A. The workflow describes a measure of tumor fraction based on ichorCNA and the limit of detection (LOD) by the SPOT-MAS test. B. Graphs show the correlation between the spike-in percentage and tumor fraction estimation in breast cancer (red), colorectal cancer (blue), gastric cancer (green), liver cancer (purple) and lung cancer (orange). Correlation was assessed by Pearson coefficient R C. Sigmoid curve shows limit of detection of 5 cancer types. LOD values are defined as as the TF levels at which the probability of detecting a cancer signal is at least 50% while maintaining at least 98% specificity (red dotted line). D. The LOD values are based on tumor fraction of five cancer types. Error bars denote 95% confidence intervals.

We next performed the SPOT-MAS test on 350 spike-in cfDNA samples and 63 pooled cfDNA samples from healthy individuals. We found that 63/63 (100%) of healthy samples were called correctly as non-cancer by the SPOT-MAS test, 129/350 (36.86%) were detected as cancer samples including 43/84 (51.19%) colorectal, 43/70 (61.43%) liver, 25/63 (39.68%) lung, 6/42 (14.29%) gastric and 12/91 (13.19%) breast cancer (**Table S5**). We next determined the LOD value for each cancer type by defining the TF at which at least 50% of cancer samples were correctly detected by SPOT-MAS while maintaining a 98% specificity (**Figure 2C**). The overall LOD for detecting all cancer samples was 0.049 (95% CI 0.043-0.059, **Figure 2D**). The lowest LOD was determined for lung cancer (0.038, 95% CI 0.027-0.098), followed by liver (0.049, 95% CI 0.043-0.059), colorectal (0.051, 95% CI 0.045-0.060), breast (0.052, 95% CI 0.041-0.18), and gastric (0.081, 95% CI 0.055-0.120) (**Figure 2D**). Our data suggested that SPOT-MAS could detect at least 50% of cancer samples at a specificity of 98% if the samples have TF of greater than 0.049.

### The impact of potential interferents on the performance of SPOT-MAS

It has been well established that blood hemolysis causes genomic DNA (gDNA) and hemoglobin contamination which could interfere with the performance of liquid biopsy tests^26^. To assess the impact of these two potential interferents, we generated 30 pooled plasma samples, including cancer (N=20) and healthy control (N=10). We then spiked these pooled plasma samples with five different concentrations of gDNA or hemoglobin before conducting SPOT-MAS tests (**Figure 3A**). All healthy control samples were correctly detected across different levels of spiked-in gDNA (**Figure 3B**) or hemoglobin (**Figure 3C**). In contrast, the false negative rates increased from 13.3% to 26.7% when the samples were contaminated with a gDNA concentration higher than 100% (**Figure 3D, Table S6**). Likewise, the false negative rates increased by 6.7% when the levels of hemoglobin contamination exceeded 500 mg/dL (**Figure 3E**, **Table S7**). Thus, our findings indicate that although gDNA and hemoglobin contamination could affect the accuracy of SPOT-MAS performance, our test remained robust at gDNA levels <100% and hemoglobin levels <500 mg/dL.

**Figure 3.**
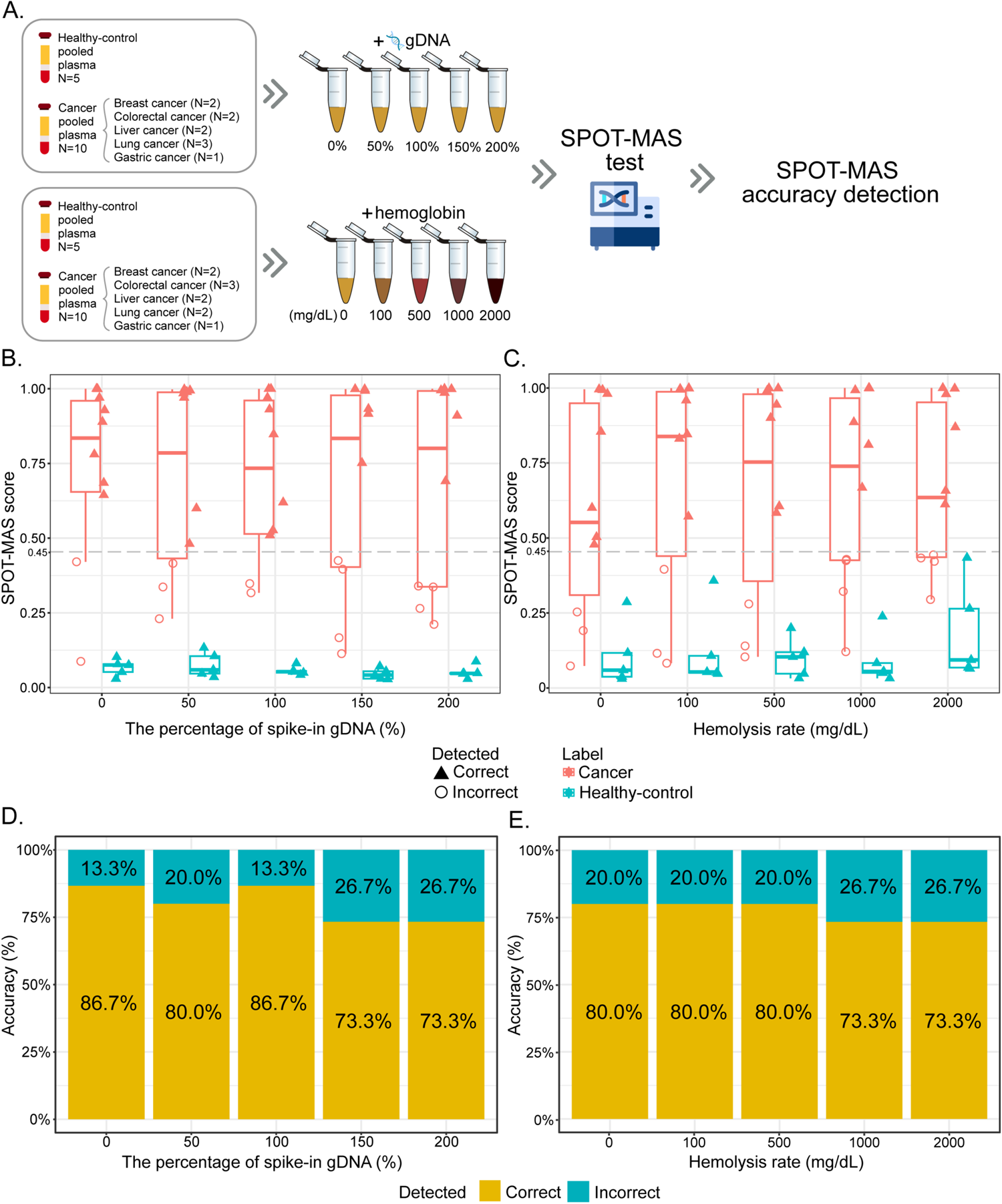
Effect of potential interferents on the performance of SPOT-MAS test. A. The workflow describes the preparation of potential interferent studies. Healthy-control plasma samples and cancer plasma samples were spiked with genomic DNA (0-200% of total cfDNA extracted from unspiked samples) and hemoglobin (0-2000 mg/dL). All samples were then subjected to the SPOT-MAS test to assess the accuracy of detection. B-C. Box plots show SPOT-MAS scores in cancer (red) and healthy-control (cyan) samples spiked with 5 differents concentrations of genomic DNA (gDNA) (B) and hemoglobin (C). Samples called as correct detected or incorrect detected are indicated by triangles or open circles, respectively. D-E. Charts show the accuracy (%) in detecting cancer and healthy-control samples spiked with 5 differents concentrations of genomic DNA (B) or hemoglobin (C).

### The repeatability and reproducibility of SPOT-MAS

To examine the repeatability and reproducibility of SPOT-MAS, we used a total of 56 pooled plasma samples (24 healthy controls and 32 cancers) and divided them into three different batches (**Figure 4A**). For the analysis of intra-batch variations, 24/24 (100%) healthy samples and 30/32 (93.75%) cancer samples were correctly detected by SPOT-MAS test across three different replicates (A, B and C); one breast cancer and one gastric cancer sample were incorrectly detected as healthy in replicate B and C, respectively (**Figure 4B, Table S8**). For the evaluation of inter-batch variations, we also observed similar levels of consistency, with 54 out of 56 (96.43%) samples being correctly detected. There was an incorrect detection in one breast sample and one gastric sample in batch 3 (**Figure 4C, Table S8**). These findings showed a highly consistent performance of SPOT-MAS assay both within runs and between runs.

**Figure 4.**
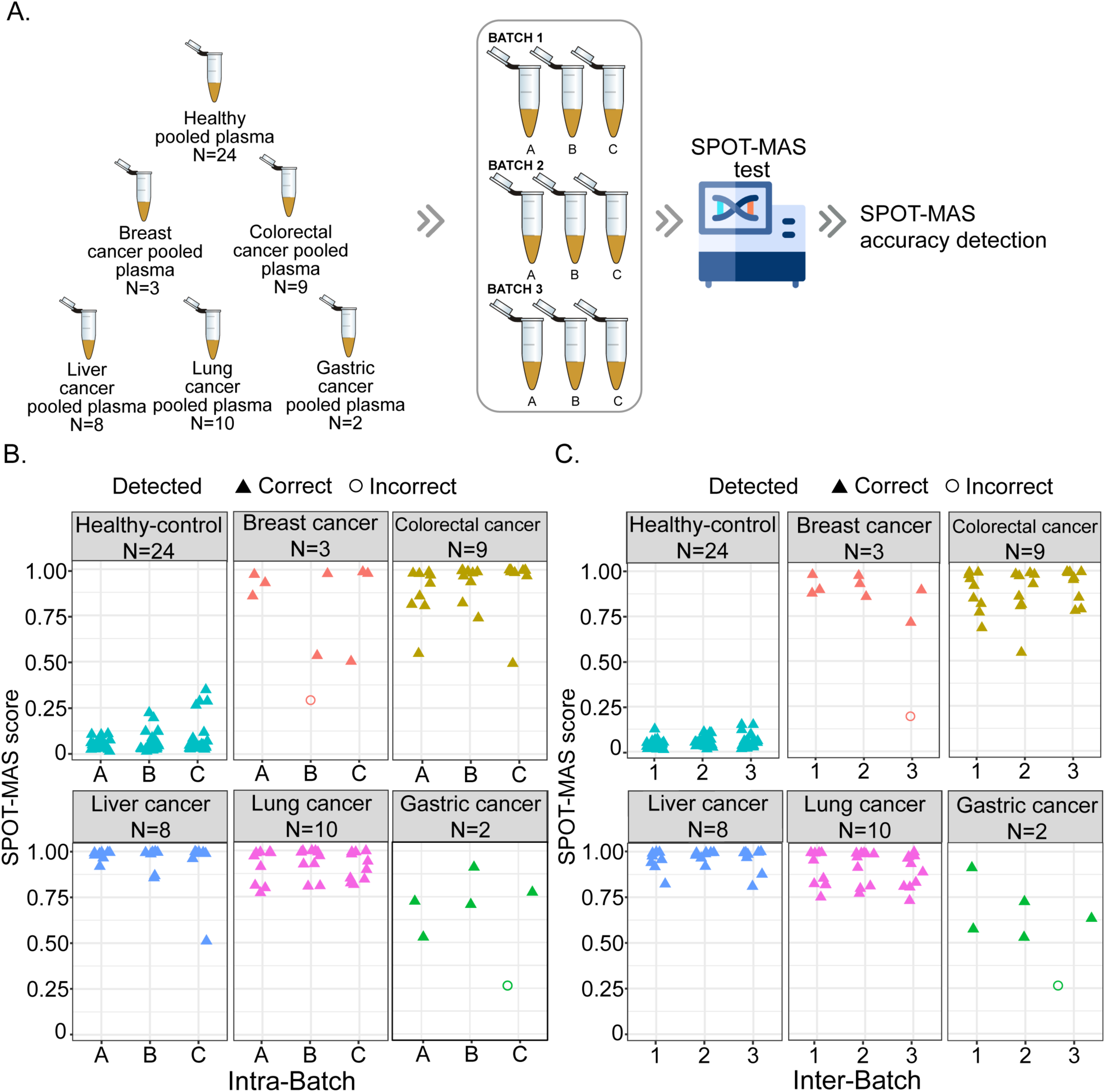
Reproducibility of SPOT-MAS test. A. The workflow shows experimental design of repeatability and reproducibility study. B-C. SPOT-MAS scores from intra-batch (B) and inter-batch (C) for breast (n=3), colorectal (n=9), gastric (n=1), liver (n=4), lung cancer (n=10), and healthy control (n=24). Samples called as correct detected or incorrect detected are indicated by triangles or open circles, respectively.

### Evaluation of clinical performance of SPOT-MAS test

Among the 9,057 eligible participants enrolled in K-DETEK, 9,024 (99.64%) completed the 12-month follow-up and were included in the study analysis, while 33 participants (0.36%) were excluded (**Table S2 and Figure 5**). The reasons for exclusion included a diagnosis of cancer (n = 1), pregnancy (n = 1), high levels of blood hemolysis (n = 31) (**Figure 5**). The clinical characteristics of the participants are summarized in **Table 1**, with a higher percentage of females than males (54.67% versus 45.33%, **Table 1**) and a median age of 50Dyears, ranging from 40 to 79 years. High-risk individuals who harbor the following risk factors including heavy smoking (16.25%), alcohol consumption (16.10%), hepatitis B/C infection (16.38%), FDR (First-Degree Relatives) diagnosed with two cancer types at an age younger than 45 or being identified as mutant carriers, (14.96%), and type 2 diabetes (5.73%) accounted for 42.27% (nD=D3,814) of all participants. The remaining 57.73% (nD=D5,210) individuals were considered moderate risk (**Table 1**). The distribution of risk factors, such as heavy smoking, hepatitis B/C infection, and type 2 diabetes was consistent with previous reports, suggesting that the K-DETEK cohort is representative of the screening population in Vietnam^27–30^.

**Figure 5.**
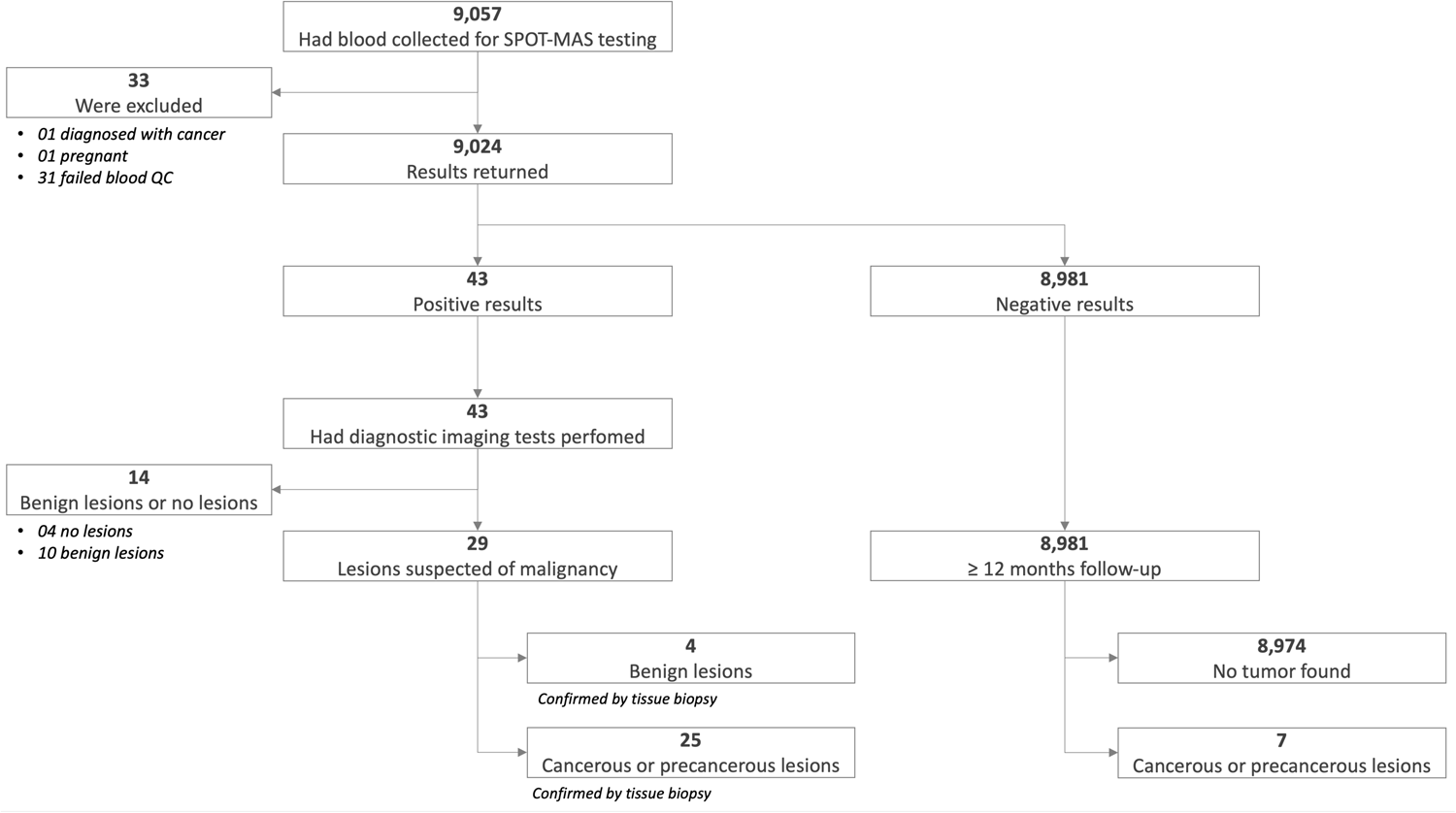
The flow chart of recruiting and following-up participants in the K-DETEK study.

**Table 1.** Clinical characteristics of 9,024 eligible participants.

| Risk classification |  | N=9024 | % |
| --- | --- | --- | --- |
| High risk |  | 3814 | 42.27 |
| Moderate risk |  | 5210 | 57.73 |
| Age | Median | 50 |  |
|  | Min | 40 |  |
|  | Max | 79 |  |
| Gender | Female | 4933 | 54.67 |
|  | Male | 4091 | 45.33 |
| Liver infection (HBV/HCV) | Yes | 1478 | 16.38 |
|  | No | 7546 | 83.62 |
| Alcohol consumptions | Yes | 1453 | 16.10 |
|  | No | 7571 | 83.90 |
| Heavy smoking | Yes | 1466 | 16.25 |
|  | No | 7558 | 83.75 |
| Diabetes | Yes | 517 | 5.73 |
|  | No | 8507 | 94.27 |
| Family history of cancer* | Yes | 1350 | 14.96 |
|  | No | 7674 | 85.04 |

\* Participants who have first-degree relatives (FDR) diagnosed with two cancer types at an age younger than 45 or being identified as mutant carriers.

The majority of participants (8,981/9,024; 99.52%) showed “ctDNA signal not detected” results and 8,974 (99.92%) of them were confirmed to be cancer-free at 12 months after enrollment, indicating a NPV of 99.92% (95%CI: 99.84-99.96) and a specificity of 99.80% (95%CI: 99.68-99.87) (**Table 2 and Figure 5**). Among these participants, 7 (0.08%) cases were found to develop cancer during the 12-month follow-up (**Table S9**). Specifically, 2 cases developed metastatic lung cancer (n=2, patients K1452 and K7249), 2 cases had locally advanced colorectal cancer (n= 2, patients K6250 and K6956), and 3 cases developed localized cancer including colon (patient K3947), lung (patient K4047) and gastric cancer (patient K6690).

**Table 2.**
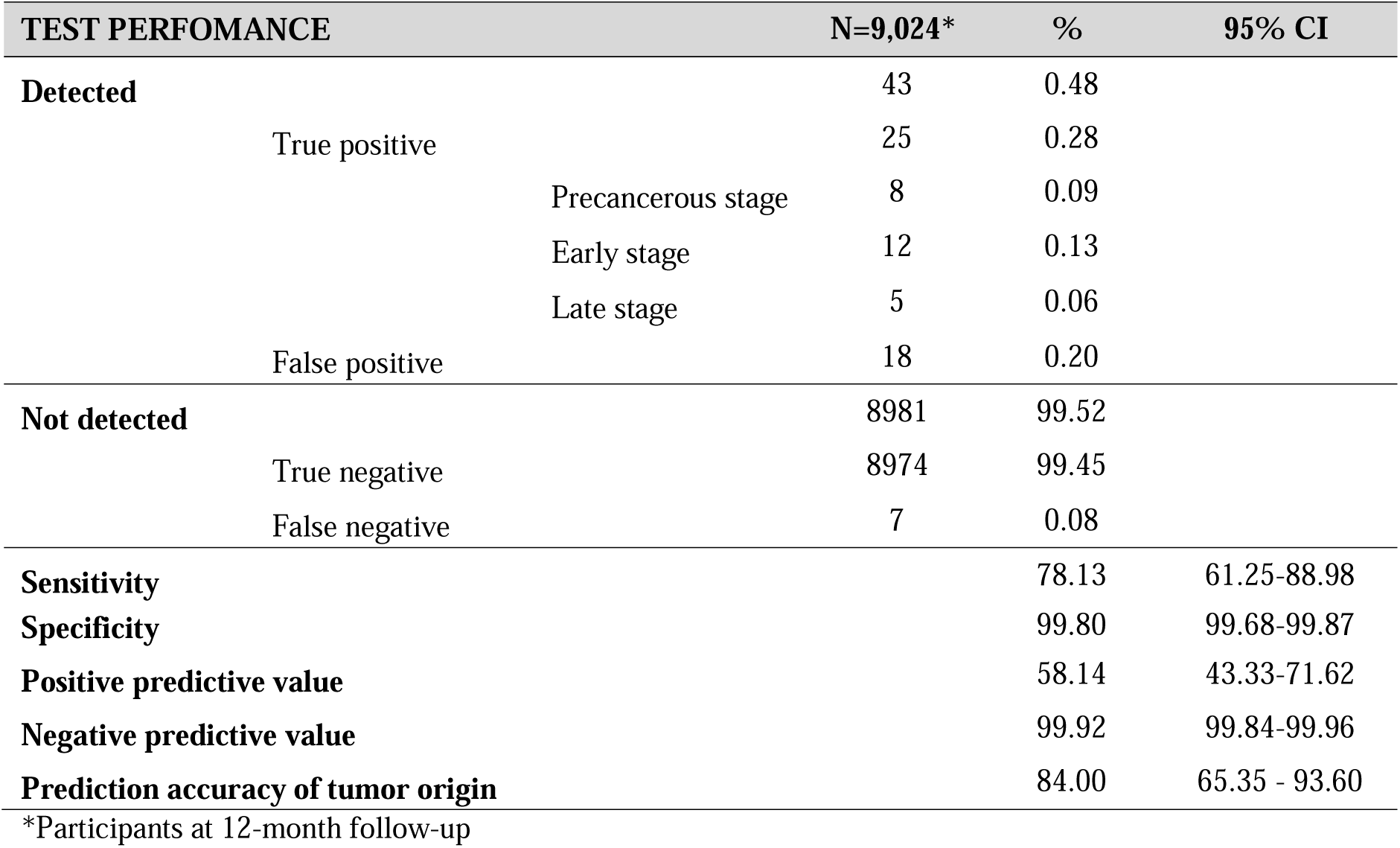
SPOT-MAS test performance.

We detected 43 cases (0.48%, **Table 2 and Figure 5**) with “ctDNA signal detected” results, all of whom were referred to undertake on-site standard of care (SOC) imaging tests to confirm the presence of tumors according to our consultation protocol (**Table S3**). All cases agreed to undertake diagnostic tests for the cancer types corresponding to the prediction of TOO provided in the SPOT-MAS test reports. Among the 43 participants with confirmed diagnostic results, 29 had imaging results with lesions suspected of malignancy and were advised to perform tissue biopsies (**Table 2 and Figure 5**). Of those, 8 (0.09%) were found to have premalignant lesions in the colon, classified under the dysplastic group (**Table 2**, **Figure 6 and Table S10**). It has been reported that individuals with these precancerous lesions are relevant for screening due to their higher risk of developing colorectal cancer compared to the general population, and early detection provides the opportunity for cancer prevention^31,32^.

**Figure 6.**
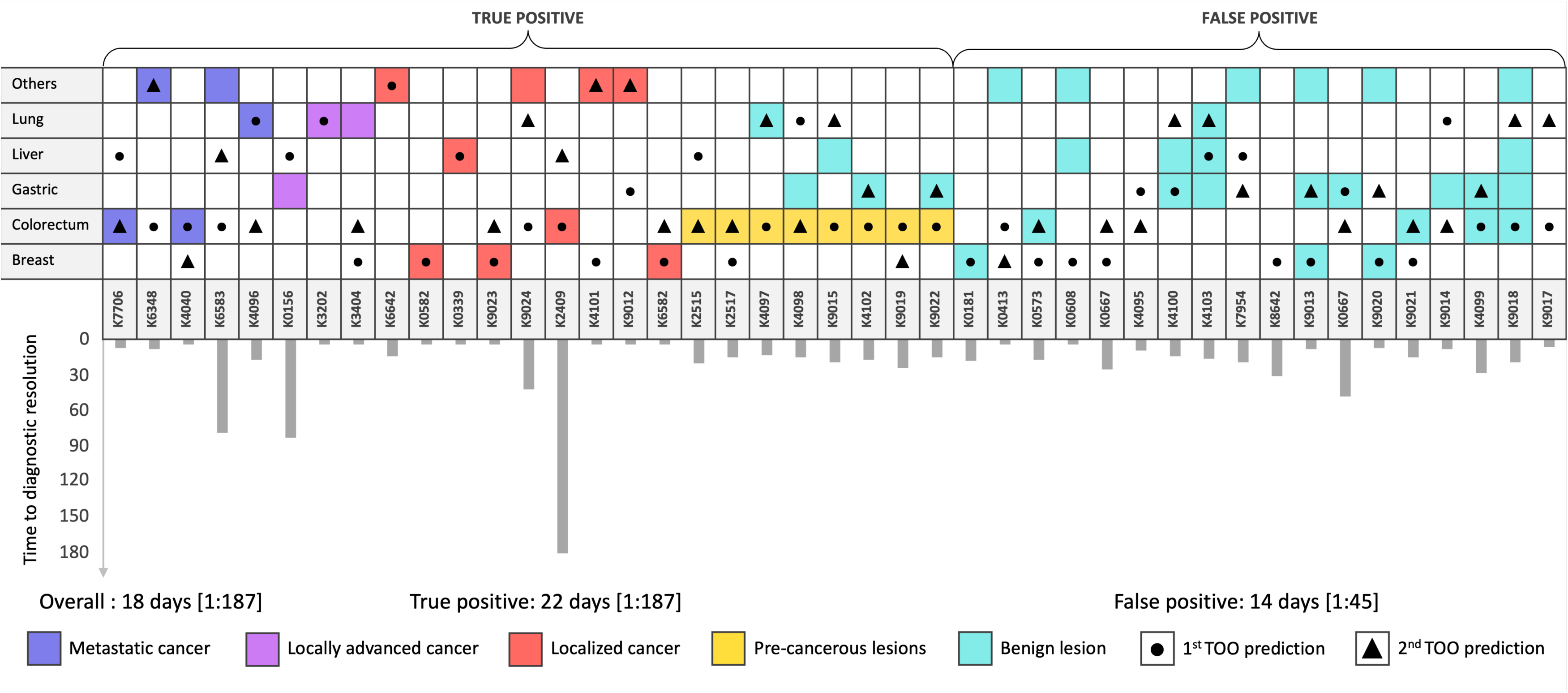
The analysis of diagnostic results of 43 participants with a ctDNA signal detected result. Colored squares show the lesion-specific origin of 43 cases with a ctDNA signal detected result, while circles and triangles denote the 1^st^ and 2^nd^ TOO prediction by SPOT-MAS. Colors indicate the cancer diagnostic outcomes, including metastatic cancer (red), locally advanced cancer (pink), localized cancer (orange), pre-cancerous lesions (yellow), and benign lesions (green). The intersections between colored squares and circles or triangles indicate the correct prediction of TOO by SPOT-MAS. The bar charts show the observed time from receipt of positive ctDNA results to final diagnosis confirmation.

Therefore, we considered these cases as true positives. In addition to precancerous cases, 12 cases (0.13%, **Table 2**) with ctDNA signal detection were diagnosed with early or nonmetastatic stage cancer (localized or locally advanced), while 5 cases (0.06%, **Table 2**) had metastatic late-stage cancer. Overall, out of the 43 cases with ctDNA signal detection, 25 were shown to have precancerous lesions or invasive cancers (true positives), suggesting a positive predictive value (PPV) of 58.14% (95% CI: 43.33-71.62) and an overall sensitivity of 78.13% (95% CI: 61.25-88.98, **Table 2**). Moreover, 21 of the 25 true positive cases developed cancer in the organs matched with either the first or second cancer type predicted by SPOT-MAS, suggesting an overall accuracy of 84.00% (95% CI: 65.35-93.60) for TOO (**Figure 6** and **Table S10**). The remaining 18 cases (0.20%, **Table 2**) with diagnostic results unable to confirm the presence of malignant or precancerous tumors were regarded as false positive cases (**Table S11**).

We observed that the median time from receipt of positive SPOT-MAS results to final diagnosis confirmation was 18 days ranging from 1 to 187 days for all 43 cases with ctDNA signal detected. Interestingly, the false positive group had a shorter median time (14 days) compared with the true positive group, which required 22 days to achieve diagnostic resolution (**Figure 6**).

Since previous studies have reported that the performance of a MCED could be dependent on the risk of target populations ^33^, we next examined such association in our cohort. We did not observe any noticeable difference in NPV and specificity across diverse groups of participants (NPV > 99.80%, specificity > 99.70% **Figure 7** and **Table S12**). By contrast, the PPV slightly increased from 57.58% (95%CI: 40.81-72.76, **Figure 7** and **Table S12**) in moderate risk participants to 60.00% (95%CI: 31.27-83.18, **Figure 7** and **Table S12**) in high-risk participants. Moreover, we observed higher PPV in the group over 50 years old as compared to the younger group < 50 years old (61.11%, 95%CI 44.86-75.22 versus 42.86%, 95% CI: 15.82-74.95, **Figure 7** and **Table S12**). For gender, the PPV was higher in male participants than female participants (80.95%, 95%CI: 60.00-92.33 versus 36.36%, 95%CI: 19.73-57.05, **Figure 7** and **Table S12**). The test exhibited a slightly lower sensitivity for early and non-metastatic stage cancer compared to metastatic late-stage cancer (70.59%, 95% CI: 46.87-86.72 versus 71.43%, 95% CI: 35.89-91.78, **Figure 7** and **Table S12**).

**Figure 7.**
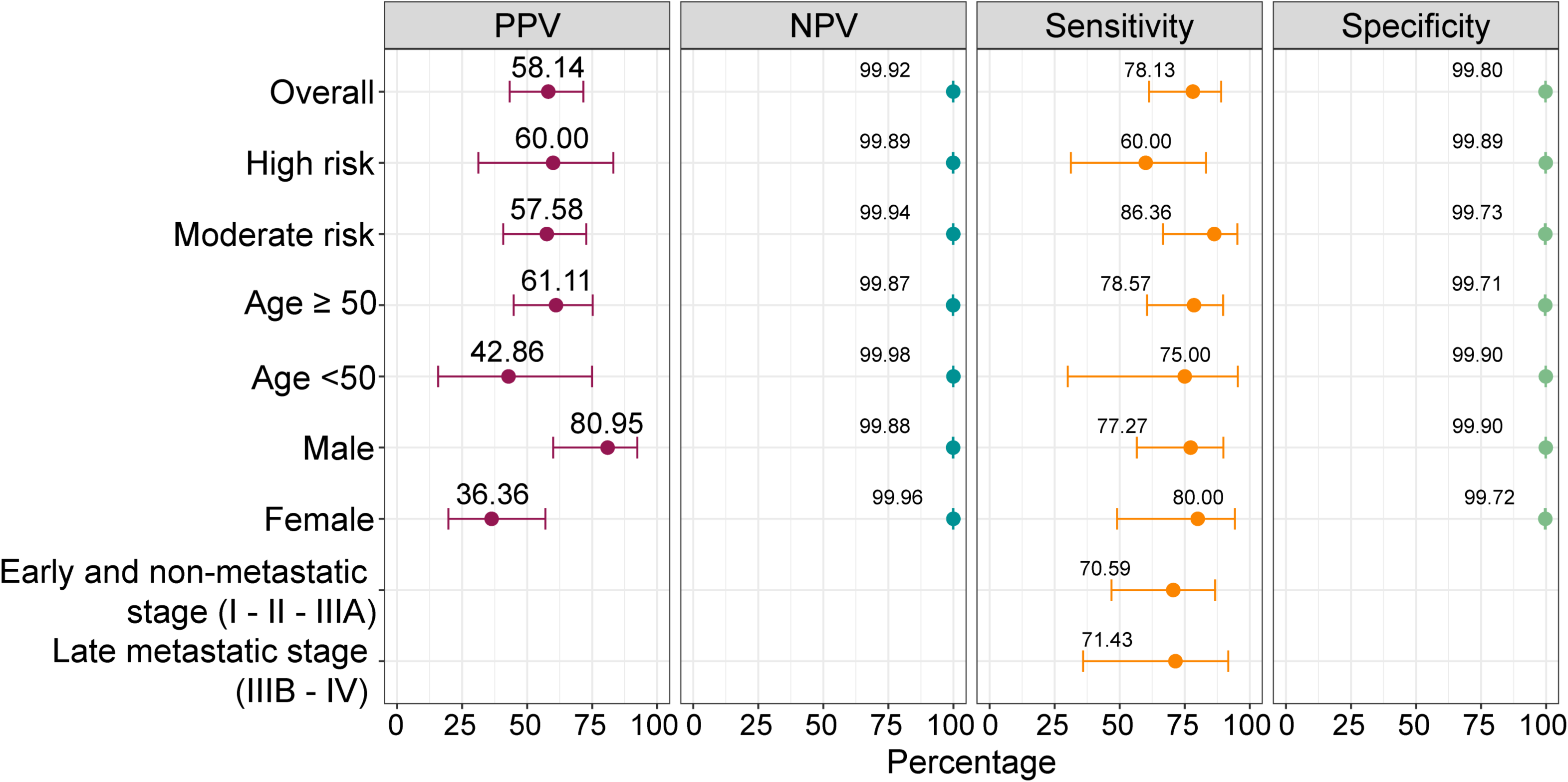
Positive predictive value (PPV), negative predictive value (NPV), sensitivity and specificity of SPOT-MAS test in different demographic groups.

Overall, our findings demonstrate that the SPOT-MAS test effectively identifies various cancer types at either pre-cancerous or early stages, exhibiting a PPV of 58.14% (95% CI: 43.33-71.62), an NPV of 99.92% (95% CI: 99.84-99.96), a sensitivity of 78.13% (95% CI: 61.25-88.98), and a specificity of 99.80% (95% CI: 99.68-99.87). In addition to its cancer detection capabilities, SPOT-MAS accurately localizes tissue-specific cancer signals, achieving an accuracy of 84.00% (95% CI: 65.35-93.60). This capability holds promise for guiding clinicians in selecting appropriate diagnostic tests for their patients.

## Discussion

The paradigm of cancer diagnosis is undergoing a significant shift with the development of MCED tests. MCED in a single blood draw test is key to successful treatment and improved survival outcomes for cancer patients. To ensure the effectiveness and reliability of MCED tests in clinical practice, thorough validation is crucial. Here, we conducted analytical validation to determine the clinical LOD and evaluate the impact of various technical factors on the robustness and accuracy of our MCED test, named SPOT-MAS. Moreover, we presented the performance of the SPOT-MAS test on a large-scale cohort of 9,024 asymptomatic participants with 12-month follow-ups.

To determine the LOD for SPOT-MAS test, we applied ichorCNA software to estimate TF of pooled cfDNA samples isolated from patients with five types of cancer. We next generated a series of cfDNA standards with different TF levels by spiking cancer cfDNA into healthy cfDNA. Of these cancer types, SPOT-MAS demonstrated the lowest LOD value for detecting lung cancer (median LOD 0.038), while showing the highest LOD value for detecting breast (median LOD 0.052) and gastric cancer (median LOD 0.081). The observed low sensitivity in detecting ctDNA signals in breast and gastric samples aligns with findings from our prior study, which validated the performance of SPOT-MAS in these sample types^21^. Moreover, our results are consistent with previous studies showing low levels of ctDNA shedding by early-stage gastric and breast tumors^10,17^.

Our previous findings showed the relationship between high hemolysis rate and its effect on isolated cfDNA and sequencing quality^26^. Therefore, we evaluated their potential interference on our assay performance in this study. Although SPOT-MAS test remained robust at hemoglobin contamination of 500 mg/dl or gDNA contamination of 100%, its performance was reduced when the amounts of contaminants were greater than these threshold values (**Figure 3B**). Our findings are in good agreement with previous studies indicating that gDNA displayed methylation and fragment profile distinct from ctDNA^34–36^, potentially introducing confounding signals to our test. Therefore, the quality of blood samples should be meticulously controlled to ensure the robustness of the test.

The SPOT-MAS workflow was built based on genome-wide low-pass sequencing to maximize its cost effectiveness^21^. However, the utilization of low sequencing depth may introduce greater variation compared to high sequencing depth, potentially diminishing the reproducibility of the test^37,38^. We therefore designed the experiment to examine the repeatability and reproducibility of SPOT-MAS. Our results demonstrated that the SPOT-MAS assay exhibited high consistency for the same samples tested either within the same sequencing runs (intra-batch) or across different sequencing runs (inter-batch) (**Figure 4**). This characteristic could be attributed to the design of our assay, which simultaneously interrogated multiple features of cfDNA to achieve a final ctDNA probability score, thus alleviating the variability that might be associated with a particular feature^21^.

The K-DETEK study was designed to determine the clinical performance of SPOT-MAS in a large asymptomatic cohort in Vietnam. The primary endpoints included the report of NPV, PPV, sensitivity, and specificity, while the secondary endpoint involved reporting TOO accuracy (**Figure 1**). Our data observed a NPV of 99.92% (95%CI: 99.84-99.96), corresponding to 8,974 out of 8,981 negative cases that remained cancer-free at 12 months after enrollment (**Table 2**). However, there were 7 participants who developed cancer within the 12-month follow-up period (false negative cases). Further analysis by ichorCNA showed that none of the 7 false negative cases displayed TF above the overall LOD of our test for detecting 5 cancer types (**Table S9 and Figure S2**). This finding indicated that all false negative cases in our study had low amounts of ctDNA in plasma beyond the capacity of SPOT-MAS test to detect. Among these cases, 2 out of 7 (patient K1452 and K7249) developed metastatic cancer. Metastatic tumors are known to display methylation signatures that differ from those of primary early-stage tumors^39^. This variation may explain why our algorithms, trained on samples meeting stringent selection criteria for early and nonmetastatic cancer (stage I-II and IIIA), did not identify these two cases.

Our findings from the K-DETEK study showed that SPOT-MAS test achieved a PPV of 58.14% (95%CI: 43.33-71.62, **Table 2**) and an overall sensitivity of 78.13% (95% CI: 61.25-88.98) for cases with precancerous lesions or invasive tumors at all stages (**Table 2**). Among the 25 true positive cases, 20 (80%) were correctly identified by SPOT-MAS test, including 8 precancerous cases and 12 early or non-metastatic tumor cases (**Table 2**). This highlights the effectiveness of SPOT-MAS in detecting precancerous lesions as well as early-stage clinical cancers. Considering precancerous cases as true positives is relevant for screening populations, as their identification allows for active prevention before they progress to invasive cancer (American Association for Cancer Research. AACR Cancer Progress Report - 2023). When considering only invasive cancers as true positives, SPOT-MAS achieved a lower PPV of 39.53% (95% CI: 26.37-54.42, **Table S13**) and higher false positive rate of 0.29% (**Table S13**). However, the performance of the SPOT-MAS test remains more advantageous than existing single-cancer screening tests with typical PPV of 4.4%-6.9% (4.4% for mammography; 6.2% for endoscopy; and 6.9% for low-dose CT scan)^40–42^ and false-positive rates of 5% to 15% per screening episode^43^. A low false positive rate is believed to reduce the number of individuals without cancer who are referred for cancer investigations, thereby directly impacting resource allocation and costs.

Eighteen cases had a ctDNA signal detected, but their diagnostic tests could not confirm the presence of a tumor (**Table 2**). While classified as false positives, it is conceivable that these cases could harbor early-stage tumors with small sizes undetectable by standard of care imaging tests. Alternatively, their tumor types might not align with the predictions of SPOT-MAS, or they could be associated with unique pathological conditions at the time of SPOT-MAS testing, leading to the generation of ‘pseudo signal’^44^. Hence, longer-term follow-up (more than 12 months) is necessary to confirm their cancer status.

It only took 18 days after receiving SPOT-MAS results to achieve diagnostic resolution (**Figure 6**). Such a short period of time for diagnostic resolution could significantly alleviate patients’ anxiety and expedite necessary interventions for those diagnosed with precancerous conditions. However, one patient (K2409) experienced the longest diagnostic resolution time of 187 days. When the patient received the SPOT-MAS test result, the patient did not exhibit any symptoms indicative of cancer and lived in a rural province in Vietnam where advanced imaging diagnostic tests were not available. Consequently, the patient chose not to undergo the recommended colon endoscopy test based on his TOO prediction by the SPOT-MAS test. After 6 months, the patient began experiencing clinical symptoms of colon cancer, including rapid weight loss and bloody diarrhea. Subsequently, a colonoscopy was performed, leading to the detection of cancer (**Table S10**). This highlights the importance of a post-test consultation procedure when applying the SPOT-MAS test in clinical practice.

Our stratification analysis showed higher PPV in older age (>= 50), male patients, or those belonging to high-risk group (**Figure 7**). This finding underscores the importance of considering the clinical characteristics and demographics of screening populations when evaluating test performance and outcomes. Our analysis of participants’ demographic characteristics (**Table 1**) closely mirrors the distribution of risk factors in the general population of Vietnam, as reported in previous studies, including 19.8% alcohol consumption^27^, 22.5% smoking^28^, 10.5% hepatitis B infection^29^, and 5.4% diabetes^30^. These similarities suggest that our study cohort is representative of the screening population in Vietnam, indicating that the test may have the potential to achieve equivalent performance when applied in clinical practice in Vietnam.

In order to compare K-DETEK findings with other clinical studies, it is essential to consider the variations in population risk and their implications on test performance^15,16^. Our K-DETEK study revealed a positive rate of 0.28% (25 true positive in 9,024 cases), higher than the cancer incident rate in Vietnamese population (0.15%)^15^. It is worth noting that the incident rate was estimated using all cancer types but with detection methods much different from our SPOT-MAS test. Our study focused on moderate- and high-risk participants with an elevated chance of developing cancer. This resulted in a PPV for detecting invasive cancer, comparable to that observed in PATHFINDER, which evaluated the multi-cancer early detection (MCED) test, named Galleri in a similar risk population and achieved a PPV of 38%^45^. Noticeably, our study detected a wide range of solid tumors and precancerous lesions, whereas the majority of cancer patients identified in PATHFINDER had hematologic cancers (48.57%). The multimodal approach of SPOT-MAS offers a comprehensive analysis through the integration of methylation, fragment length profile, DNA copy number aberration and end motif in a single library reaction. This unique approach could explain the differences observed in comparison to the Galleri test used in the PATHFINDER study, which primarily focused on methylation makers^21,46^.

Our study has several limitations. First, the determination of limit of detection by ichorCNA tool, which relies on CNA signals, might not provide an accurate reflection of the true LOD of SPOT-MAS test, particularly for the cancer types with low CNA signals. Second, our assay has lower sensitivity for certain cancer types with low shedding tumors, such as gastric and breast, or metastatic tumors, resulting in false negative cases. Moreover, SPOT-MAS only focuses on five common cancer types, potentially missing rarer cancer types. To further enhance the sensitivity of early cancer detection and provide a broader spectrum of cancer types, future studies are needed to explore the multi-omics approach that combine different makers, such as cfDNA, cfRNA and circulating tumor cells^47^. Third, the number of true positive cases (17 cases, 0.19%) with confirmed malignant lesions is relatively small due to the rigorous selection of patients without cancer-related symptoms. Thus, our future clinical trial is ongoing to evaluate the performance of the SPOT-MAS test in symptomatic participants. Finally, it remains unclear whether a MECD test such as SPOT-MAS could enhance survival. Future randomized trial studies are needed to address this question. These studies will provide valuable insights into the effectiveness of the test and its impact on patient outcomes.

## CONCLUSION

In conclusion, our study demonstrated that SPOT-MAS is a promising multi-cancer blood test for early cancer detection. Through comprehensive analytical validation, we have established the robustness of SPOT-MAS test, with a limit of detection of 0.049 (95% CI: 0.043-0.059) tumor fraction. Importantly, the data from K-DETEK study, the largest prospective trial in Asia, highlights the potential utility of SPOT-MAS in limited-resource countries. Together, our study offers compelling evidence for the feasibility of SPOT-MAS in early detection of asymptomatic cancer patients, thereby enhancing overall population health outcomes.

## Data Availability

The analytical data are available upon reasonable request by email to the corresponding authors (LST, MDP). Raw sequencing data are not publicly available due to ethical and regulatory restrictions.

## Declarations

## Ethics approval and consent to participate

This study was approved by the Ethics Committee of the Medic Medical Center, University of Medicine and Pharmacy and Medical Genetics Institute, Ho Chi Minh city, Vietnam. Written informed consent was obtained from each participant in accordance with the Declaration of Helsinki.

## Availability of data and materials

The analytical and clinical data are available upon reasonable request by email to the corresponding authors (LST, MDP). Raw sequencing data are not publicly available due to ethical and regulatory restrictions.

## Author contribution

Formal analysis: Luu Hong Dang Nguyen, Thi Hue Hanh Nguyen, Ngoc Minh Phan, Ba Linh Tieu, Dac Ho Vo, Trung Hieu Tran, Thanh Dat Nguyen, Van Thien Chi Nguyen, Y-Thanh Lu, Trong Hieu Nguyen, Vu Uyen Tran, Minh Phong Le, Thi Minh Thu Tran, Minh Nguyen Nguyen, Thi Tuong Vi Van, Anh Nhu Nguyen, Thi Thanh Nguyen, Huu Tam Phuc Nguyen,, Chi Thuy Tien Cao

Patient consultancy and screening: Luu Hong Dang Nguyen, Thi Hue Hanh Nguyen, Van Hoi Le, Vinh Quang Bui, Lan Hieu Nguyen, Nhu Hiep Pham, Thanh Hai Phan, Huu Thinh Nguyen, Van Song Tran, Chi Viet Bui, Van Kha Vo, Pham Thanh Nhan Nguyen, Ha Huu Phuoc Dang, Van Dung Pham, Van Thinh Cao, Ba Linh Tieu, Thi Thanh Nguyen, Huu Tam Phuc Nguyen, Chi Thuy Tien Cao, Van Tung Nguyen, Thi Le Quyen Le, Thi Lan-Anh Luong, Thi Kim Phuong Doan, Thi Trang Dao, Canh Duy Phan, Thanh Xuan Nguyen, Nguyen Tuong Pham, Bao Toan Nguyen, Thi Thu Thuy Pham, Huu Linh Le, Cong Thanh Truong, Thanh Xuan Jasmine, Minh Chi Le, Van Bau Phan, Quang Binh Truong, Thi Huong Ly Tran, Minh Thien Huynh, Tu Quy Tran, Si Tuan Nguyen, Vu Tran, Van Khanh Tran, Huu Nguyen Nguyen, Duy Sinh Nguyen, Thi Van Phan, Thi Thanh-Thuy Do, Dinh Kiet Truong, Hung Sang Tang

Data Curation: Luu Hong Dang Nguyen, Thi Hue Hanh Nguyen, Nhu Nhat Tan Doan, Hoang Tan Nguyen, Phuoc Loc Doan, Le Anh Khoa Huynh, Tien Anh Nguyen

Methodology: Duy Sinh Nguyen, Thi Van Phan, Thi Thanh-Thuy Do, Dinh Kiet Truong, Hung Sang Tang, Minh-Duy Phan, Hoa Giang, Hoai-Nghia Nguyen, Le Son Tran

Conceptualization: Huu Nguyen Nguyen, Duy Sinh Nguyen, Thi Van Phan, Thi Thanh-Thuy Do, Dinh Kiet Truong, Hung Sang Tang, Minh-Duy Phan, Hoa Giang, Hoai-Nghia Nguyen, Le Son Tran

Writing-original draft: Luu Hong Dang Nguyen, Thi Hue Hanh Nguyen, Giang Thi Huong Nguyen, Le Son Tran

Writing-Review and Editing: Giang Thi Huong Nguyen, Minh-Duy Phan, Hoa Giang, Hoai-Nghia Nguyen, Le Son Tran

## Acknowledgments

We thank all participants who agreed to take part in this study, and all the clinics and hospitals who assisted in patient consultation and sample collection. We thank Gia Bao Nguyen, George School, PA, USA, for his help in proofreading this manuscript.

---

**Figure S1.**
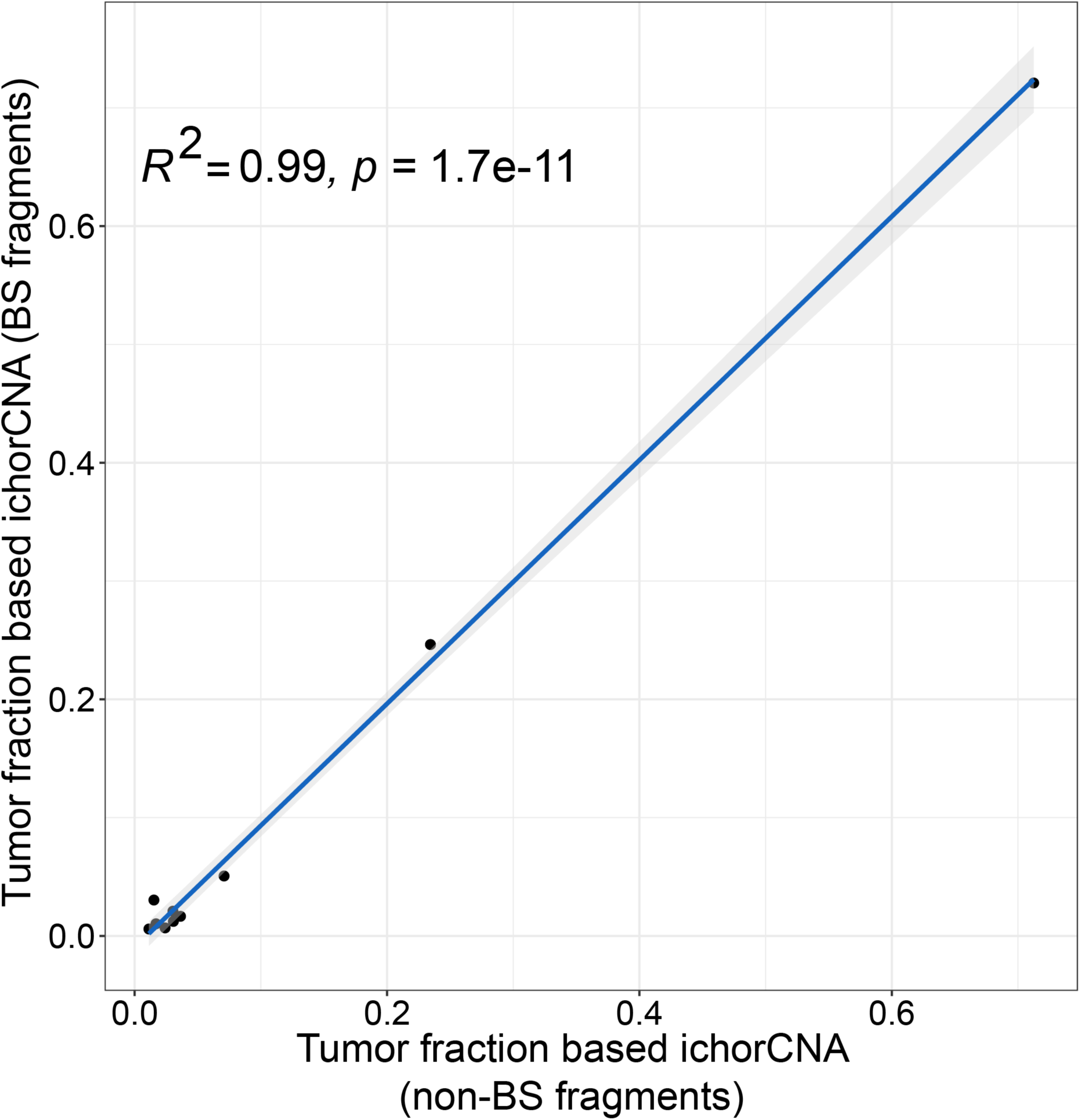
Correlation of tumor fraction by ichorCNA between non-bisulfite fragments and bisulfite treated fragments.

**Figure S2.**
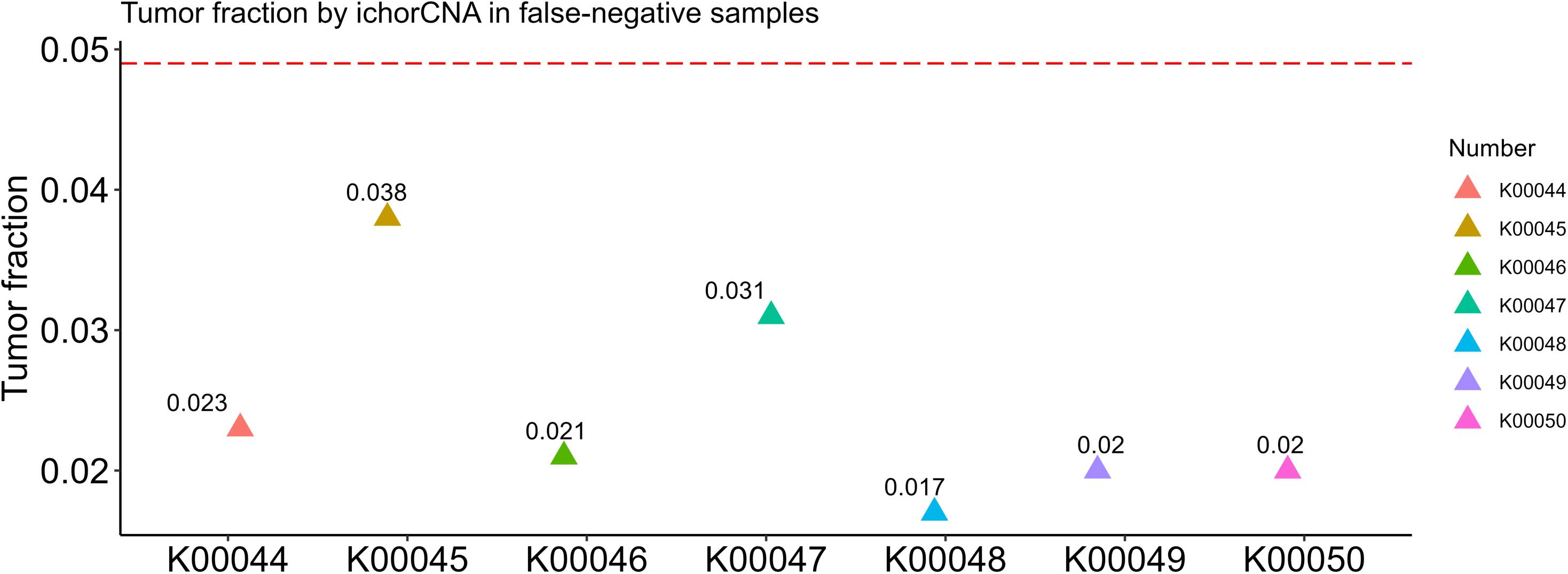
Tumor fraction by ichorCNA in false negative samples.

Table S1: Demographic details of samples used for analytical validation

Table S2: Demographic details of eligible 9,024 participants enrolled in K-DETEK study

Table S3: Standard of care imaging tests used for diagnostic resolution

Table S4: Tumor fraction of healthy control and spike-in cancer cfDNA determined by ichorCNA and SPOT-MAS score

Table S5: Summary of SPOT-MAS detection results of spike-in cfDNA samples from 5 cancer types

Table S6: SPOT-MAS results of gDNA spiked-in samples

Table S7: SPOT-MAS results of hemoglobin spiked-in samples

Table S8: SPOT-MAS results of samples in reproducibility experiment

Table S9: Standard of care imaging test and tumor fraction by ichor CNA results of false negative cases

Table S10: Standard of care imaging test results of true positive cases

Table S11: Standard of care imaging test results of false positive cases

Table S12: Sensitivity, specificity, positive predictive value, negative predictive value of SPOT-MAS test in different demographic groups

Table S13: SPOT-MAS test performance without precancerous lesions

